# Blood Flow Restriction Training After ACL Reconstruction: A Systematic Review of Early-Phase strength, morphological, pain, and neuromuscular adaptations

**DOI:** 10.1101/2025.10.12.25337802

**Authors:** Abdelmounaim Assermouh, Rita Atik, Sonia Redouane

**Affiliations:** Universidad Europea – Real Madrid Graduate School, Madrid, Spain

**Keywords:** ACLR, blood flow restriction, low-load resistance training, strength, pain, hypertrophy, neuromuscular adaptation

## Abstract

**Objective:** This systematic review aimed to evaluate the effects of blood flow restriction (BFR) training compared to traditional rehabilitation or high-load resistance training (HL-RT) on quadriceps strength, muscle morphology by cross-sectional area (CSA), pain and neuromuscular activation (NMA) following anterior cruciate ligament reconstruction (ACLR).

**Methods:** A systematic review was performed according to PRISMA 2020 guidelines. Five databases (PubMed, Web of Science, MEDLINE Complete, CINAHL, and SPORTDiscus) were searched using the following equation : (blood flow restriction training OR blood flow restriction exercise OR blood flow restriction therapy OR BFR therapy OR KAATSU training OR occlusion resistance training) AND (anterior cruciate ligament injury OR ACL injury OR anterior cruciate ligament tear OR ACL tear OR ACL reconstruction). Only randomized controlled trials (RCTs) were included. Methodological quality and risk of bias were assessed using the Cochrane Risk of Bias tool and the PEDro scale.

**Results:** Eight RCTs (n = 234) met the inclusion criteria. BFR training enhanced quadriceps strength and preserved or increased muscle CSA compared with low-load rehabilitation, while reducing pain and improving electromyographic activity. Outcomes were directionally consistent and comparable to HL-RT but achieved under substantially lower mechanical stress. Heterogeneity was moderate, primarily related to occlusion pressure calibration and session dosage. No serious adverse events were reported.

**Conclusion:** BFR training appears to be a safe and effective adjunct for early ACLR rehabilitation, enhancing quadriceps strength and morphology while reducing pain under low mechanical loads. Its effects are comparable to HL-RT protocols, with emerging evidence of neuromuscular benefits. Further standardization of pressure and dosage parameters is needed to refine its clinical application.

## Introduction

Although surgical techniques and postoperative protocols have evolved substantially, persistent quadriceps deficits remain among the most clinically relevant sequelae of ACLR (Krishnan & Williams, 2011). These chronic impairments—characterized by reductions in strength, CSA, and NMA—can persist for months or even years after surgery (Lepley, 2015; Palmieri-Smith et al., 2008). They extend beyond mechanical weakness, disrupting knee kinetics, altering motor control strategies, and increasing the risk of re-injury or early-onset osteoarthritis (Buckthorpe et al., 2019; Hart et al., 2015).

Beyond peripheral deficits, growing evidence highlights substantial neuroplastic alterations following ACL injury and reconstruction (Neto et al., 2019).

Neuroimaging studies demonstrate decreased activation of primary motor and somatosensory cortices with a compensatory over-reliance on visual and prefrontal regions during movement tasks (D. Grooms et al., 2015; D. R. Grooms et al., 2015). These cortical reorganizations reflect disrupted sensorimotor integration and reduced automaticity of movement (Strong et al., 2022). Consequently, rehabilitation after ACLR should aim not only to restore muscular strength and morphology but also to recalibrate cortical and sensorimotor control to reestablish efficient and automatic motor patterns (Calabrò et al., 2025).

Together, these peripheral and central alterations underline the need for rehabilitation strategies that target both muscular recovery and neural reorganization to optimize long-term knee function and performance. Rehabilitation following ACLR therefore aims to restore the mechanical and neural components of function to ensure a safe and effective return to sport (RTS) (Kotsifaki et al., 2025).

HLRT, typically prescribed at 65–80% of one-repetition maximum (1RM), remains the gold standard for promoting hypertrophy and strength adaptation (Schoenfeld et al., 2017). However, in the early postoperative phase, HL-RT may be contraindicated due to graft vulnerability, pain, effusion, or concomitant meniscal lesions (Kacprzak, 2025). This creates a clinical need for interventions capable of inducing both muscle and neural adaptations under reduced mechanical stress.

Originally developed in Japan as KAATSU training in the 1960s, BFR training has gained attention as a novel adjunct to rehabilitation (Freitas et al., 2021). By applying a pneumatic cuff to partially occlude venous return while maintaining arterial inflow, BFR induces local hypoxia and metabolic stress that stimulate anabolic signaling pathways (mTORC1, GH, IGF1) and promote recruitment of type II muscle fibers even at loads as low as 20–30% 1RM (Loenneke, Wilson, et al., 2012; Pearson & Hussain, 2015; Scott et al., 2016). In addition to peripheral hypertrophy, BFR has been shown to enhance motor-unit recruitment and EMG amplitude, suggesting potential neuromuscular and cortical benefits during early-phase ACLR rehabilitation (Hughes, Rosenblatt, et al., 2019b; Jung et al., 2022).

Recent RCTs integrating BFR into ACLR rehabilitation have reported improvements in quadriceps strength, CSA preservation, and pain modulation without compromising graft safety (Hughes, Patterson, et al., 2019a; Ohta et al., 2003; Vieira de Melo et al., 2022).

Nevertheless, inconsistencies in occlusion pressure, cuff width, intervention duration, and training parameters have produced heterogeneous outcomes across studies. With the publication of several high-quality RCTs in recent years, an updated synthesis is warranted to clarify the physiological, neuromuscular, and clinical effects of BFR training in ACLR rehabilitation.

Therefore, the aim of this systematic review was to evaluate the effects of BFR training compared with conventional rehabilitation following ACLR, focusing on its impact on muscle hypertrophy , strength, pain, and NMA.

## Methods

### Study Design and Registration

This review followed 2020 PRISMA guidelines (Page et al., 2021). The protocol defined the eligibility criteria, search strategy, and analysis framework a priori and was prospectively registered in the PROSPERO database (registration number: CRD420251165961; available at https://www.crd.york.ac.uk/prospero/display_record.php?ID=CRD420251165961).

The primary objective was to evaluate the effects of BFR training, compared with conventional rehabilitation or HL-RT, on quadriceps strength, muscle morphology, pain, and NMA after ACLR. *Eligibility criteria*

Studies were selected according to the PICOS framework:

- Population: adults or adolescents undergoing postoperative rehabilitation after primary ACLR, with no restriction on sex or graft type.
- Intervention: exercise-based rehabilitation incorporating BFR training during the postoperative period.
- Comparison: conventional rehabilitation without BFR or HL-RT protocols.
- Outcomes: quadriceps strength, muscle CSA or thickness, pain, and NMA measured through surface electromyography (EMG).
- Study design: randomized controlled trials (RCTs).

Exclusion criteria included non-randomized or observational studies, animal or in vitro models, BFR interventions applied pre- or perioperatively, studies comparing different BFR modalities, and articles without full-text availability or quantitative outcome data. Publications not written in English were excluded.

### Search Strategy

The sources of information were obtained by the research of studies that was carried out using PubMed, Web of Science, MEDLINE Complete, CINAHL with full text, and SPORTDiscus databases. The terms used for the literature search were: (blood flow restriction training OR blood flow restriction exercise OR blood flow restriction therapy OR BFR therapy OR KAATSU training OR occlusion resistance training) AND (anterior cruciate ligament injury OR ACL injury OR anterior cruciate ligament tear OR ACL tear OR ACL reconstruction). The final search was conducted on 12 september 2025. Two independent reviewers (A.A. and R.A.) conducted the search using the same methodology, and the differences were resolved by consensus. The reference sections of the original studies were screened manually.

### Study selection and Data Extraction

All references were imported into Mendeley Reference Manager (Elsevier, New York, USA) for citation management. Duplicates were automatically removed and verified manually.

Two reviewers independently screened titles and abstracts to exclude irrelevant papers, followed by full-text review to confirm eligibility. Discrepancies were resolved by consensus.

Data extracted from each study included authorship, year of publication, sample size, participant characteristics, graft type, timing of intervention, occlusion method (absolute mmHg or % limb occlusion pressure—LOP/AOP), training load (%1RM), frequency, duration, exercise type, and reported outcomes (strength, CSA, pain, EMG). Extraction accuracy was verified by a second reviewer.

### Quality assessment

Methodological quality and risk of bias were assessed using the Cochrane Risk of Bias 2 tool and the Physiotherapy Evidence Database (PEDro) scale. Each study was evaluated across domains of randomization, allocation concealment, blinding, completeness of data, and selective reporting.

PEDro scores were used to classify trials as poor (≤3), fair (4–5), good (6–8), or excellent (≥9) quality. Disagreements between reviewers were resolved through discussion and consensus.

### Data Synthesis

Because of methodological and clinical heterogeneity across the included RCTs—particularly in occlusion pressures, training loads, intervention duration, and outcome measures—a quantitative meta-analysis was not feasible.

A narrative synthesis was therefore conducted, organized by outcome domain: (1) muscle strength, (2) muscle morphology , (3) pain, and (4) NMA. Directional consistency of effects (positive, neutral, or negative) was analyzed qualitatively. Statistical heterogeneity (I²) was not calculated due to incomplete effect size reporting.

Where available, findings were interpreted considering clinically meaningful thresholds such as the minimal detectable change (MDC) rather than isolated statistical significance.

## Results

### Study selection

The database search identified 710 records. After removal of 212 duplicates, 498 titles and abstracts were screened.

Following full-text assessment of 21 potentially eligible articles, eight randomized controlled trials (RCTs) met the inclusion criteria (Curley et al., 2021; Curran et al., 2020; DE MELO et al., 2022; Hughes, Patterson, et al., 2019b; Hughes, Rosenblatt, et al., 2019a; Iversen et al., 2016; Jung et al., 2022; Ohta et al., 2003).

All studies investigated postoperative BFR training in patients following ACL reconstruction (ACLR).

A PRISMA flow diagram summarizing the selection process is presented in Figure 1.

**Figure 1.**
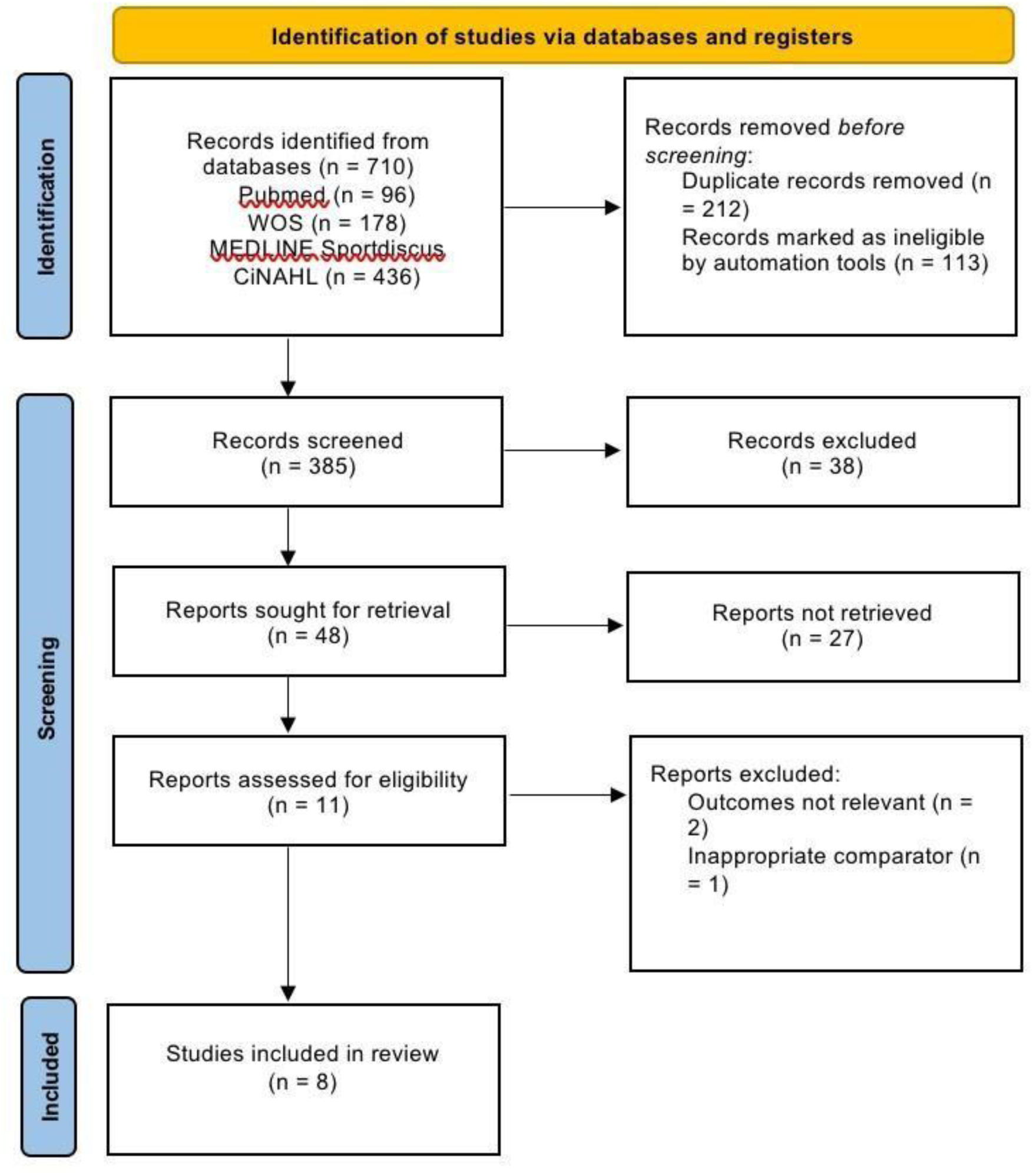
Flow diagram of literature search according to PRISMA guidelines 2020.

### Study characteristics

The eight included RCTs involved a total of 234 participants (mean age range: 19–35 years; 75% male).

Intervention duration ranged from 2 to 16 weeks, with training frequencies between 2 and 5 sessions per week.

BFR was generally applied during low-load resistance training (20–30% 1RM) with occlusion pressures between 40–80% limb occlusion pressure (LOP) or 130–240 mmHg when expressed in absolute terms.

Exercises most commonly included leg press and knee extension/flexion, usually initiated between the second and fourth postoperative week.

Comparators involved either conventional low-load rehabilitation or HL-RT, 65–80% 1RM. Primary outcomes were quadriceps strength (8/8), muscle CSA(5/8), pain (6/8), and NMA via EMG (3/8).

A detailed description of study populations, designs, outcomes, and main findings is provided in Table 1.

**Table 1.**
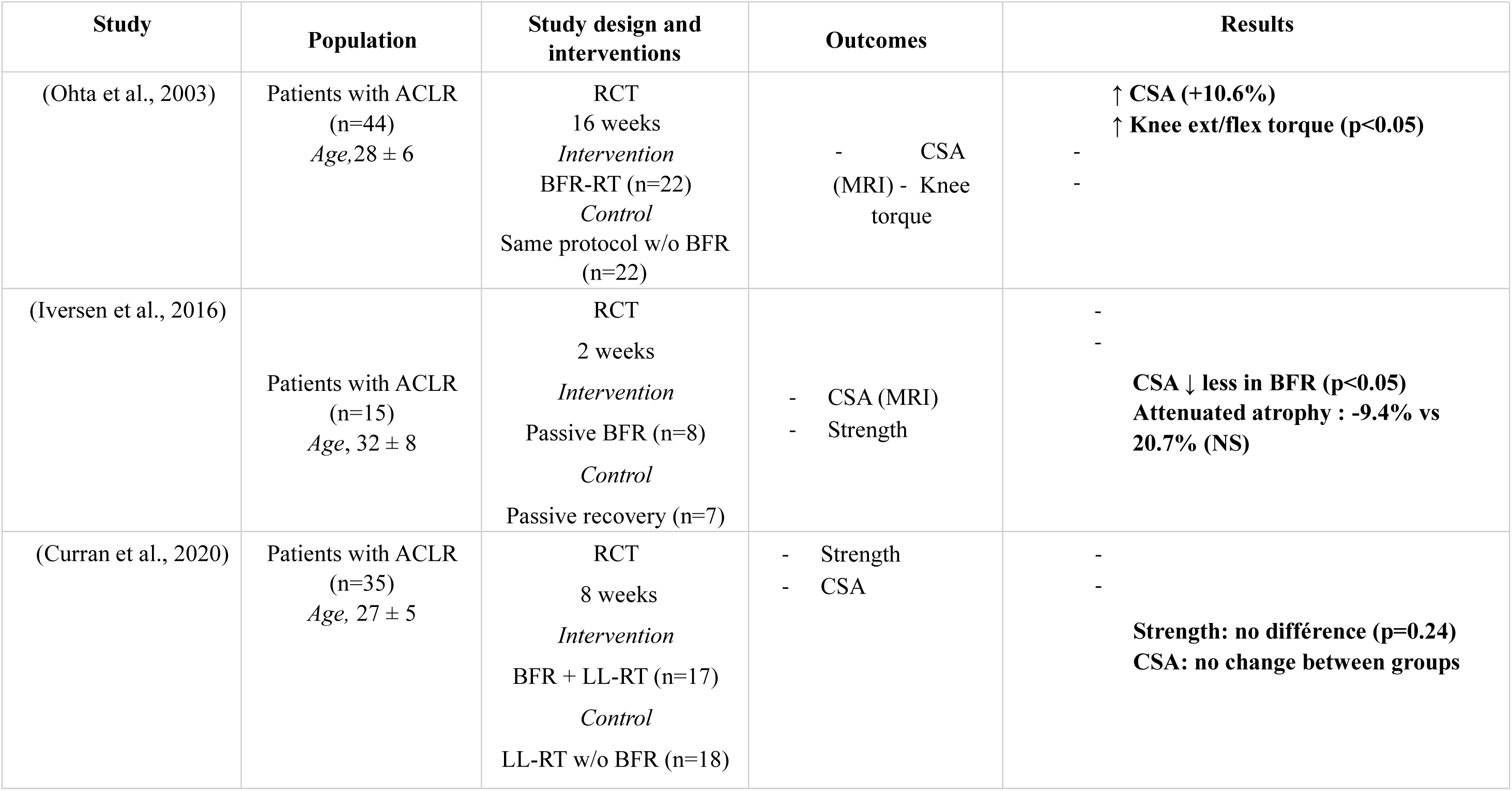
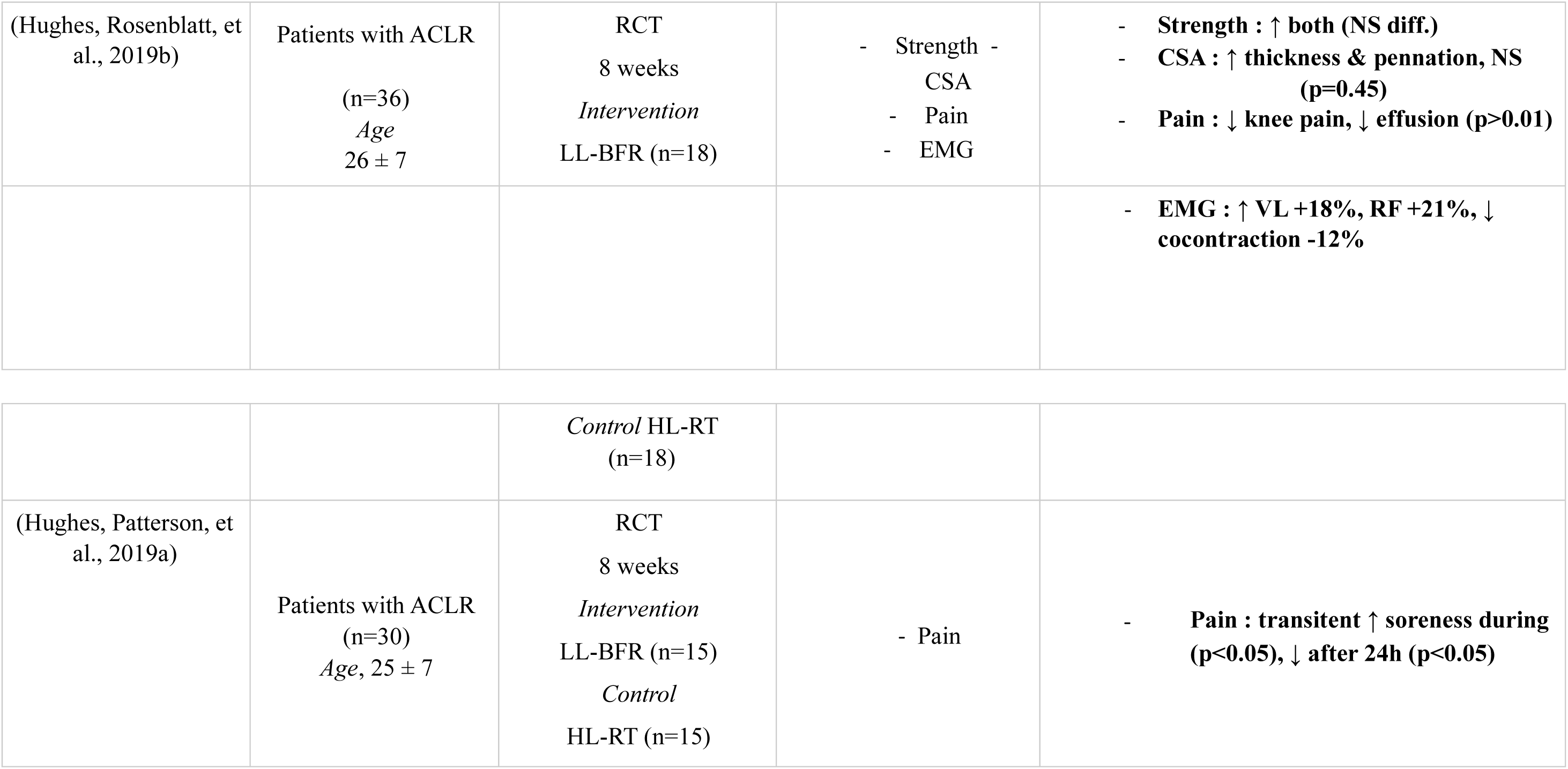
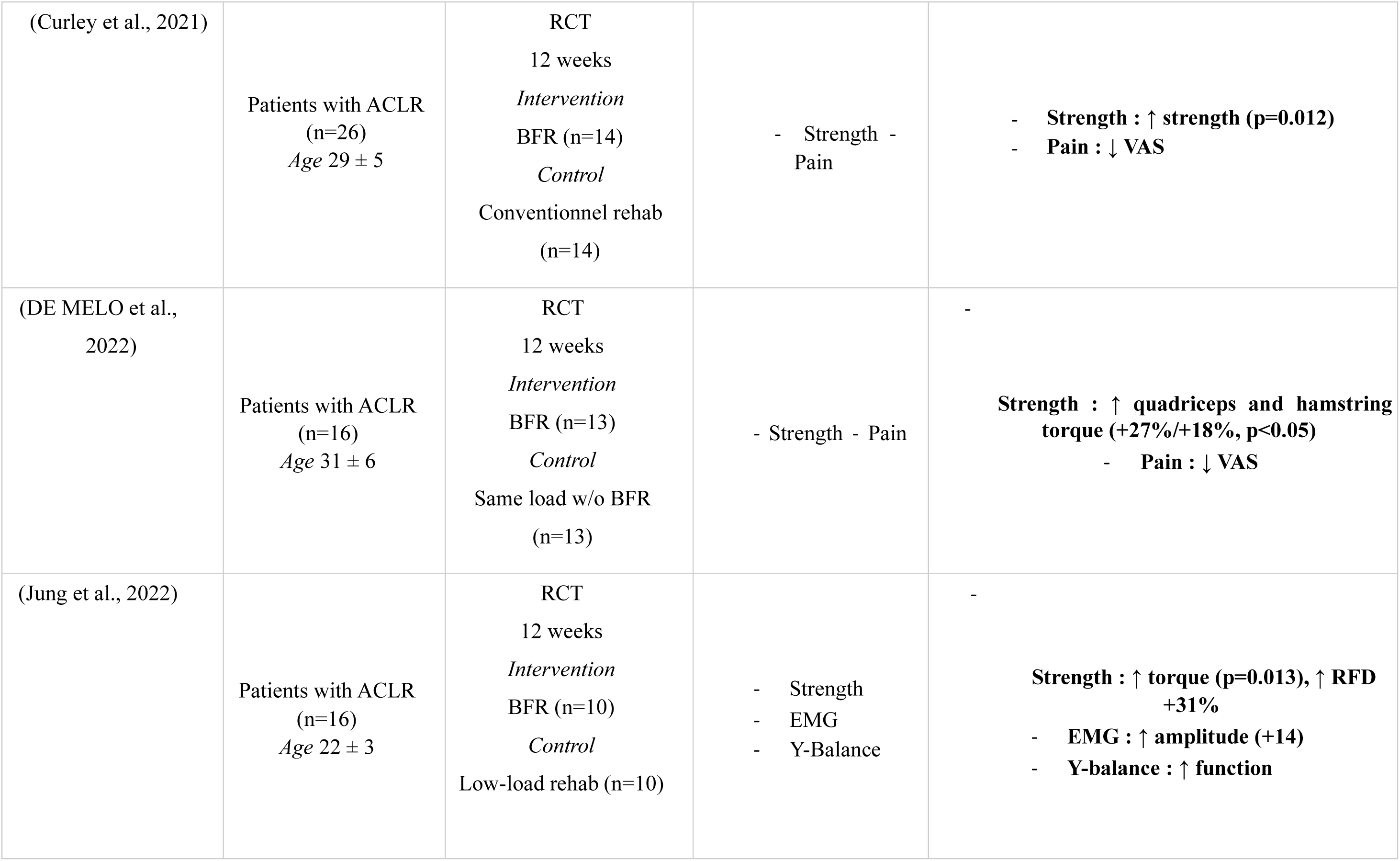
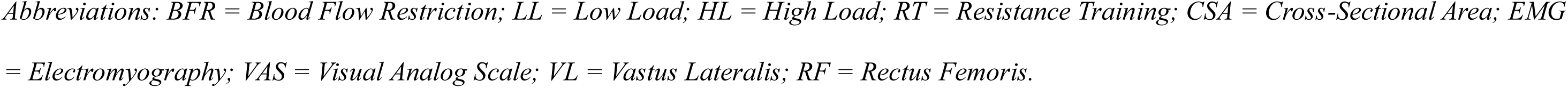
Characteristics and results of the included studies.

A summary of intervention parameters is presented in Table 2.

**Table 2.** Intervention variables extracted from each included study.

| Study | BFR Pressure | Load (%1RM) | Frequency (sessions/week) | Duration | Exercises |
| --- | --- | --- | --- | --- | --- |
| (Ohta et al., 2003) | 200 mmHg (fixed) | 30% | 2-3 | 16 weeks | Leg press Knee ext/flex |
| (Iversen et al., 2016) | 130 mmHg | Passive | 14 (2/day x 7 days) | 2 weeks | Passive occlusion |
| (Curran et al., 2020) | 80% AOP | 30% | 2-3 | 8 weeks | Leg press Knee ext/flex |
| (Hughes, Rosenblatt, et al., 2019b) | 80% LOP | 30% | 2 | 8 weeks | Leg press Knee ext |
| (Hughes, Patterson, et al., 2019a) | 80% LOP | 30% | 2 | 8 weeks | Knee ext |
| (Curley et al., 2021) | 40% LOP | 20-30% | 3 | 12 weeks | TKE<br>Squats<br>Step-ups |
| (DE MELO et al., 2022) | 40-80% LOP | 30% | 3 | 12 weeks | Leg press Knee ext/flex |
| (Jung et al., 2022) | 60% LOP | 30% | 3 | 12 weeks | Leg ext Balance task |

*Abbreviations:*
*BFR = Blood Flow Restriction; 1RM = One-Repetition Maximum; AOP = Arterial Occlusion Pressure; LOP = Limb Occlusion Pressure; ext/flex = Extension/Flexion; TKE = Terminal Knee Extension.*

### Risk of Bias and Methodological Quality

PEDro scores ranged from 6 to 8, indicating fair to good methodological quality.

All studies reported randomization, and six described adequate allocation concealment. Participant and therapist blinding were not feasible due to the nature of the intervention, representing the main risk of bias.

Outcome assessor blinding was implemented in five studies.

According to the Cochrane RoB2 tool, six studies were rated as low risk, and two as some concerns, mainly due to incomplete adherence reporting.

A detailed overview of the risk of bias is presented in Table 3, the summary of risk of bias is illustrated in Figure 2, and Table 4 reports The PEDro scale scores.

**Figure 2.**
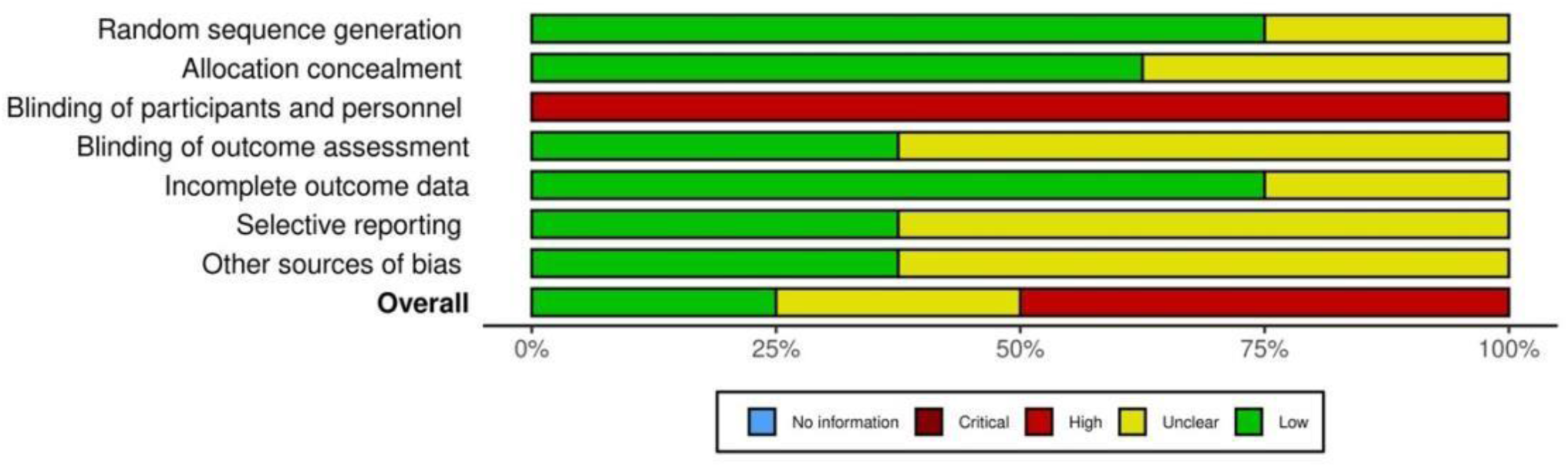
Risk of bias summary. Review authors’ judgements about each risk of bias item for each included study (Risk of Bias scale) and risk of bias graph. Review authors’ judgements about each risk of bias item presented as percentages across all included studies (Risk of Bias scale).

**Table 3.**
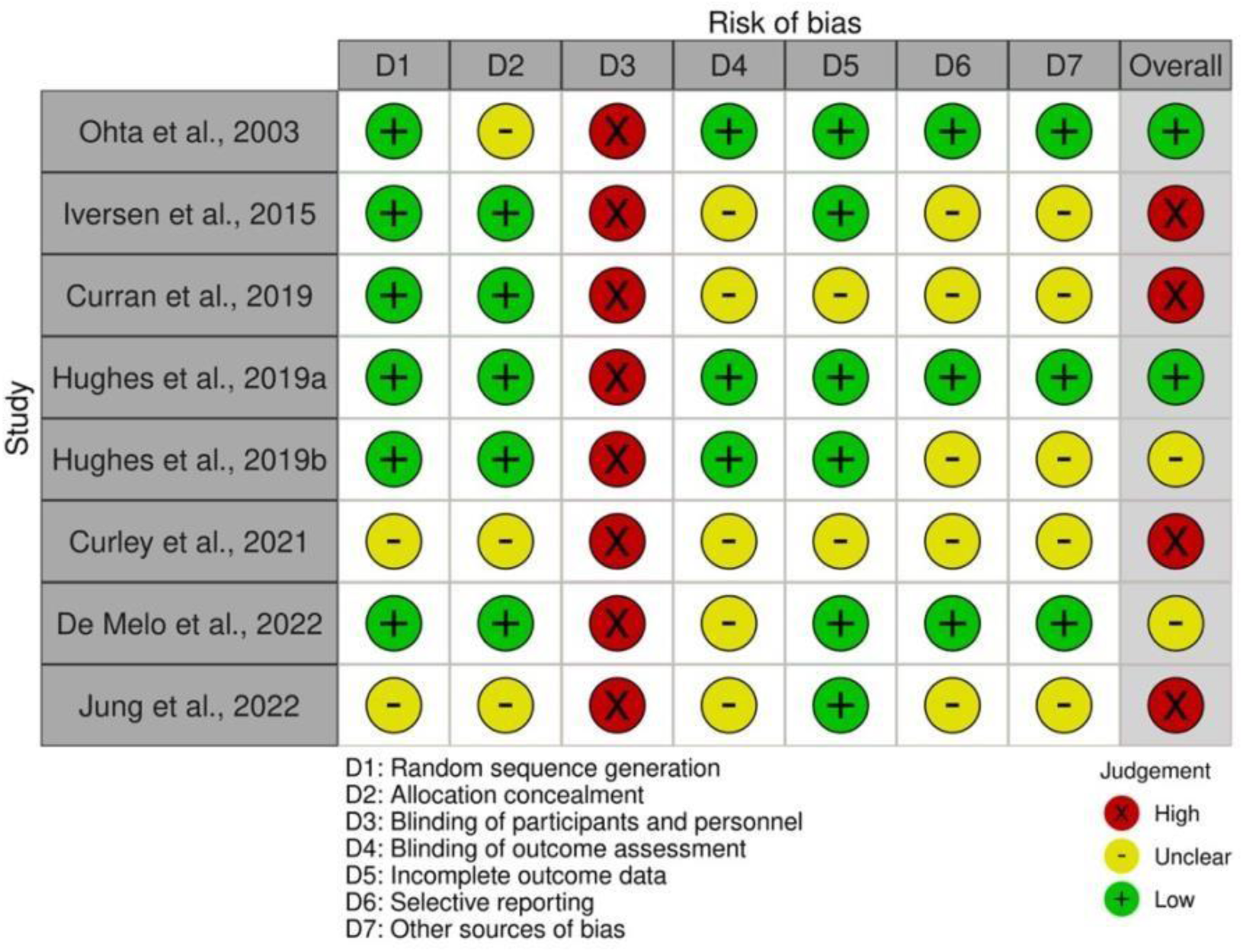
Risk of bias.

**Table 4.** Assessment of the studies’ quality based on the PEDro Scale.

|  | Items |  |  |  |  |  |  |  |  |  |  |  |
| --- | --- | --- | --- | --- | --- | --- | --- | --- | --- | --- | --- | --- |
|  | 1 | 2 | 3 | 4 | 5 | 6 | 7 | 8 | 9 | 10 | 11 | Total |
| (Ohta et al., 2003) | 1 | 1 | 0 | 1 | 0 | 0 | 1 | 1 | 0 | 1 | 1 | 7 |
| (Iversen et al., 2016) | 1 | 1 | 0 | 1 | 0 | 0 | 0 | 1 | 0 | 1 | 1 | 6 |
| (Curran et al., 2020) | 1 | 1 | 0 | 1 | 0 | 0 | 0 | 1 | 0 | 1 | 1 | 6 |
| (Hughes, Rosenblatt, et al., 2019b) | 1 | 1 | 1 | 1 | 0 | 0 | 1 | 1 | 0 | 1 | 1 | 8 |
| (Hughes, Patterson, et al., 2019a) | 1 | 1 | 1 | 1 | 0 | 0 | 1 | 1 | 0 | 1 | 1 | 8 |
| (Curley et al., 2021) | 1 | 1 | 0 | 1 | 0 | 0 | 0 | 0 | 0 | 1 | 1 | 5 |
| (Vieira de Melo et al., 2022) | 1 | 1 | 0 | 1 | 0 | 0 | 0 | 1 | 0 | 1 | 1 | 6 |
| (Jung et al., 2022) | 1 | 1 | 0 | 1 | 0 | 0 | 0 | 1 | 0 | 1 | 1 | 6 |

1, patient choice criteria are specified; 2, random assignment of patients to groups; 3, hidden assignment; 4, groups were similar at baseline; 5, all patients were blinded; 6, all therapists were blinded; 7, all evaluators were blinded; 8, measures of at least one of the key outcomes were obtained from more than 85% of baseline patients; 9, intentiontotreat analysis was performed; 10, results from statistical intergroup comparisons were reported for at least one key outcome; 11, the study provides point and variability measures for at least one key outcome

### Quadriceps strength

All eight RCTs quantified quadriceps strength via isokinetic, isometric, or 1RM testing.

Six demonstrated statistically significant or clinically relevant improvements with BFR versus conventional rehabilitation.

- (Ohta et al., 2003) – 16-week BFR program produced +20.7 ± 2.2 % increase in kneeextension torque and +18.9 ± 2.0 % in flexion versus +9.4 ± 1.6 % in controls (p < 0.05).
- (Iversen et al., 2016) – After 2 weeks of passive BFR, quadriceps atrophy was limited (–9.4 %) compared with control (–20.7 %, p < 0.05).
- (Curran et al., 2020) – No between-group difference in MVIC or 1RM (Δ ≈ 8 %, p = 0.24).
- (Hughes, Rosenblatt, et al., 2019b) – LL-BFR and HL-RT achieved similar gains (+14 % strength, p > 0.05).
- (Vieira de Melo et al., 2022) – BFR group +24 % isokinetic strength vs +10 % in LL- RT (p < 0.05).
- (Jung et al., 2022) – Peak torque +16.5 % (60°/s) and +12.3 % CSA (p < 0.05).
- (Curley et al., 2021) – Strength +10 %, significant vs baseline (p = 0.012), not between groups.
- (Hughes, Patterson, et al., 2019a) – Focused on pain; strength unchanged.

Across trials, strength gains averaged +15–30 %, consistently exceeding the minimal detectable change (∼10 %) and frequently matching HL-RT outcomes under far lower loads.

### Pain

Six RCTs evaluated pain (VAS or NRS).

Five reported significant reductions with BFR; one (Curran 2019) showed parity with HL-RT.

- (Hughes, Patterson, et al., 2019a) – VAS decrease –2.1 ± 0.7 during exercise (p < 0.01) and –1.8 ± 0.6 after 24 h (p < 0.05).
- (Vieira de Melo et al., 2022) – ≈ 35 % mean VAS reduction after 12 weeks.
- (Curley et al., 2021) – VAS drop ≈ –1.5 points (p < 0.05).
- (Iversen et al., 2016; Jung et al., 2022; Ohta et al., 2003) – reported similar decreases (p < 0.05).

Overall reduction ranged –1.2 to –2.8 points on a 10-point scale, suggesting consistent hypoalgesic effects likely mediated by altered afferent input and metabolic stress. *CSA* Five studies assessed CSA by MRI or ultrasound.

Three (Ohta 2003; De Melo 2022; Jung 2022) found significant hypertrophy or preservation with BFR:

- (Ohta et al., 2003) – CSA +10.6 % vs –2.5 % in controls (p < 0.05).
- (Iversen et al., 2016) – Atrophy –9.4 % vs –20.7 % (p < 0.05).
- (Vieira de Melo et al., 2022) – Muscle thickness +12 % vs +3 % (p < 0.05).
- (Jung et al., 2022) – CSA +12.3 %, p < 0.05.
- (Curran et al., 2020; Hughes, Rosenblatt, et al., 2019b) – no significant differences (Δ ≈ 6 %, p > 0.05).

Collectively, BFR mitigated early postoperative atrophy and promoted moderate hypertrophy compared with conventional therapy.

### NM activation EMG

Only two studies evaluated EMG activity.

- (Hughes, Rosenblatt, et al., 2019b) – Significant increases: vastus lateralis +18 %, rectus femoris +21 %, co-contraction –12 % (p < 0.05).
- (Jung et al., 2022) – Enhanced EMG amplitude across quadriceps heads (p < 0.05).

These results indicate improved neural drive and recruitment efficiency under low mechanical stress.

### Safety

No thrombotic, neurologic, or graft-related events were reported.

Minor petechiae or transient discomfort occurred in two trials and resolved spontaneously.

BFR was safe and well tolerated across all interventions.

### Descriptive Heterogeneity and Outcome Summary

Heterogeneity was moderate, mainly from occlusion-pressure calibration, cuff width, exercise selection, and rehabilitation timing (2–16 weeks post-ACLR).

Despite this, the direction of effects was consistent across trials.

A synthesis appears in Table 5.

**Table 5.** Summary of Outcomes and Directional Consistency.

| Outcome | Studies (n) | Direction (BFR vs Control) | Consistency | Magnitude/Notes |
| --- | --- | --- | --- | --- |
| **Quadriceps Strength** | 8 | 6 ↑, 2 = | High | ≈ 15–30 % gain; > MDC (≈ 10 %) |
| **Muscle CSA / Thickness** | 5 | 3 ↑, 2 = | Moderate | +10–12 %; atrophy prevention |
| **Pain** | 6 | 5 ↓, 1 = | High | –1 to –3 VAS points |
| **EMG Activation** | 2 | 2 ↑ | Moderate | ↑ VL +18 %, RF +21 %, ↓ co-contract –12 % |
| / | 8 | None Serious | High | Minor, transient |
| **Overall** |  |  | Moderate-High | Consistent direction of benefit |

### Summary Statement

Across the eight RCTs, BFR training improved quadriceps strength (≈ 15–30 %), increased or preserved CSA (+10–12 %), reduced pain (–1 to –3 VAS points), and enhanced EMG activation (+18–21 %) without adverse events.

Effects were comparable to HL-RT yet achieved under low mechanical stress, supporting BFR as a safe, clinically meaningful adjunct for early ACLR rehabilitation.

## Discussion

The primary aim of this review was to analyze the effects of BFR training on quadriceps strength, CSA, pain, and NM activation in patients following ACLR. Across the eight RCTs included, BFR generally improved quadriceps performance and muscle preservation compared with conventional low-load rehabilitation, while achieving outcomes comparable to HL-RT under substantially lower mechanical stress. These findings suggest that BFR may represent a useful adjunct during the early to mid-stages of ACLR rehabilitation, particularly when traditional loading is limited by postoperative constraints.

### Interpretation of findings

Collectively, the evidence indicates that BFR can elicit meaningful improvements in quadriceps strength and morphology, even in the presence of graft protection and restricted joint loading. The magnitude of these changes—often within the range of 15–30% for strength and 8–12% for CSA—suggests that the metabolic and neural stimuli provided by BFR are sufficient to mitigate early postoperative atrophy and functional decline. Although the precise mechanisms remain partially inferred, enhanced neural drive, increased recruitment of fast-twitch fibers, and elevated local anabolic signaling may all contribute to these adaptations (Centner & Lauber, 2020; Frechette et al., 2025; Loenneke, Fahs, et al., 2012; Wahli et al., 2024).

### Neuromuscular and functional relevance

Persistent quadriceps inhibition remains one of the major barriers to full functional recovery after ACLR. Several trials included in this review reported improvements in EMG amplitude, voluntary activation, and dynamic stability metrics following BFR application. While these findings cannot be regarded as direct evidence of cortical or spinal plasticity, they support the notion that BFR may indirectly promote the reactivation of inhibited motor pathways by alleviating pain and joint effusion and by modulating afferent input from group III–IV muscle fibers, thereby enhancing neuromuscular excitability in the early stages of rehabilitation (Liu et al., 2024). Such indirect modulation of excitability could explain the observed gains in torque production and control precision during the early rehabilitation phases (Taleshi et al., 2025).

### Clinical implications

From a clinical standpoint, BFR provides a structured approach to stimulate muscular and neuromuscular recovery when conventional loading remains unsafe or poorly tolerated. Its integration can shorten the period of low-load training, allowing earlier restoration of quadriceps capacity without compromising graft integrity (Kacprzak, 2025). When individualized based on limb occlusion pressure (40–80% LOP) and performed under professional supervision, BFR appears both feasible and safe (Hughes et al., 2018). Nonetheless, its role should be considered complementary rather than substitutive to progressive strengthening. Combining BFR with targeted neuromuscular and proprioceptive exercises may optimize long-term outcomes and prepare patients for higher-level functional tasks (Kotsifaki et al., 2023).

### Limitations and future directions

Despite consistent directionality across studies, methodological heterogeneity remains a limiting factor. Differences in cuff pressure calibration, exercise selection, and intervention duration complicate cross-study comparisons and preclude quantitative synthesis. Most trials were short-term, included small samples, and focused exclusively on early postoperative outcomes. The long-term influence of BFR on power, rate of force development (RFD), interlimb symmetry, and return-to-sport readiness therefore remains uncertain. Future trials should adopt standardized protocols, larger cohorts, and follow-up assessments extending into the later rehabilitation phases to clarify the durability and functional significance of these effects.

### Safety considerations and conclusion

When appropriately prescribed and monitored, BFR represents a safe and pragmatic addition to early ACLR rehabilitation. Across available RCTs, adverse events were rare and limited to transient discomfort, supporting its clinical feasibility under individualized occlusion parameters. Beyond its mechanical practicality, BFR introduces a structured framework for maintaining quadriceps activation during protected loading periods. However, the current body of evidence does not yet establish its long-term translational impact on function, performance, or reinjury prevention. Future research should clarify dose–response relationships, refine screening criteria, and evaluate integration within progressive, criterion-based rehabilitation pathways. Until then, BFR should be applied judiciously—as an adjunct that bridges protection and progression—rather than a replacement for evidence-based strengthening and neuromuscular retraining strategies.

## Conclusions

BFR training is a feasible and evidence-supported adjunct for early ACLR rehabilitation. It promotes quadriceps preservation, strength recovery, and pain reduction under low mechanical load, serving as a safe bridge between graft protection and functional reactivation. Evidence remains moderate due to small samples and heterogeneous protocols, highlighting the need for standardized pressure calibration and training parameters. When properly applied, BFR may complement traditional loading and help optimize early rehabilitation outcomes.

## Data Availability

All data supporting the findings of this review are contained within the published studies included in the analysis. Extracted data and summary tables are available from the corresponding author upon reasonable request.

## Data Availability

All data used in this review are available within the article and its referenced sources.

